# Modelling the long-term demographic and epidemiological trends in Malawi

**DOI:** 10.1101/2025.09.25.25336670

**Authors:** Rachel E. Murray-Watson, Margherita Molaro, Tara D. Mangal, Sakshi Mohan, Bingling She, Sangeeta Bhatia, Joseph H. Collins, Eva Janoušková, Tim Colbourn, Andrew N. Phillips, Timothy B. Hallett

## Abstract

Malawi is facing a dual burden of disease, with an increase in non-communicable diseases coinciding with still-high infectious disease burdens. What this will look like in the future, and how it will impact demand for healthcare, is unknown. In this study, we use the Thanzi La Onse (TLO) model - an individual-based “all diseases – whole health-system” model calibrated to Malawi’s demographic, epidemiological, and healthcare data - to project population, disease burdens, and healthcare demand from 2020 to 2070.

We project Malawi’s population to grow from 19.5 million in 2020 to 54.2 million in 2070, with median age rising from 17 to 25 years. Infectious disease burdens, particularly HIV/AIDS, TB, malaria, and acute respiratory infections, will decline, though cancers, cardiometabolic diseases, and mental health disorders burdens will increase and account for over one-third of disability-adjusted life years by 2070. Demand for healthcare grows across all cadres, with the steepest increases in clinical and mental health services.

Our results highlight the epidemiological and demographic shifts projected to occur in Malawi. In particular, we show the shift away from infectious and disease of childhood toward NCDs and age-related conditions will require adaptations in Malawi’s health system.

## 2 Introduction

Malawi has seen decades of sustained improvement in health outcomes. Between 2000 and 2022, neonatal and maternal mortality fell by 50% and 36% respectively [21, 20]. New HIV infections fell from a peak of 14 per 1,000 people in 1992 to 0.61 per 1,000 in 2023 [19], and life expectancy at birth has increased by over 18 years since 2000 [35]. These gains have been achieved despite the limited healthcare resources available [8].

The lowered mortality rate, in particular, has ushered in the beginnings of a demographic transition. In addition, theory suggests that the shifting mortality patterns, alongside changes in a range of socioeconomic and environmental factors, will lead to a so-called “epidemiological” transition whereby age-related diseases and lifestyle eventually supplant infectious diseases as the primary cause of morbidity and mortality [27].

Indeed, in Malawi, non-communicable diseases (NCDs) and key risk factors have increased in prevalence over the few decades, with nearly 40% of deaths attributable to NCDs in 2018 [7]. Risk factors for a range of conditions, such as hypertension [10] and adiposity [7] are also found to be prevalent in older age groups, indicating that disease incidence may worsen as the population ages.

However, this increase in NCDs accompanies rather than replaces a sustained high prevalence of infectious diseases [23, 34]. This “double burden” of diseases [22, 17] has also been described in other low- and middle-income countries [1, 4, 5, 34]. Despite a growing understanding of this double burden, comparatively little is known about what these long-term changes will demand from the healthcare system. Yet to effectively plan for evolving healthcare needs, such long-term population and epidemiological projections are essential.

Whilst some demographic and epidemiological projections for Malawi exist (e.g. [34]), they overlook the interdependence between health states and the complex interactions that shape an individual’s overall health status. Additionally, few studies have addressed what these interacting epidemiological and demographic changes will mean in terms of future demand for healthcare workers [28]. This paper aims to address this gap by utilising *Thanzi La Onse* (TLO, www.tlomodel.org, [8]), an all-disease, whole-health system model. We aim to make long-term projections of Malawi’s population and associated changes in the causes of mortality and disability-adjusted life years (DALYs), as well as project the future demand for healthcare services.

## 3 Methods

### 3.1 Model overview

The TLO model is an individual-based model that is calibrated to available demographic, epidemiological, and health system data in Malawi [8]. It captures Malawi’s changing demography and epidemiology, including the prevalence of infectious and NCDs (accounting for around 80% of the mortality and 73% of the DALYs in Malawi), and their risk factors. It also models healthcare demand and provision via the healthcare-seeking behaviour of the population and the availability and effectiveness of diagnostics and interventions. The model also tracks the availability of healthcare workers, consumables, and hospital beds, though we assume that there are no restrictions on healthcare worker time or bed availability, which could lead us to over-estimate the amount of healthcare services delivered. We do assume that there is limited diagnostic accuracy, that referrals for further treatment are not always made when required, and that treatment is limited by healthcare worker competence. The parameter values for each of these will vary depending on the disease. For example, the probability that hypertension being diagnosed is 0.38, whilst the probability of stunting being diagnosed in a generic appointment is 0.01.

Calibrated parameters determine the probability that an intervention will be delivered. For example, in delivering emergency obstetric and newborn care, the correct intervention will be delivered 60.2% of the time. A full description of the model, and of the data sources adopted in its calibration, can be found in [8, 28].

### 3.2 Lifestyle Factors

The model accounts for changes in lifestyle factors over time, which affect both a person’s likelihood of accessing healthcare and their risk of developing certain diseases. For example, if an individual initiates tobacco use, their risk of developing cardiometabolic disease increases. Conversely, if they gain access to a non-wood-burning stove, the risk decreases. The impacts of these lifestyle factors on the probability of developing certain conditions are determined using regression models. We assume that the following lifestyle factors increase in the population over time: urban living, higher body mass index (BMI), increased salt and sugar intake, reduced physical activity, tobacco use, and improved access to clean water, non-wood-burning stoves, handwashing facilities, and sanitation. For a full description of all lifestyle factors, and how they change in prevalence, see www.tlomodel.org and SI Figure 6.

We note that even though the parameter values do not change over time, changes in the prevalence of risk factors, disease, and healthcare arise endogenously in this system. For example, the prevalence of a high BMI will increase with an ageing population, though the relationship between age and BMI remains constant between 2020 and 2070.

### 3.3 Healthcare Seeking Behaviour and Healthcare Access

An individual’s probability of seeking healthcare after symptom onset is determined by a regression model that uses socio-demographic factors (such as region of residence or age) as inputs, alongside symptom severity. Certain subgroups - such as those in the lowest wealth quintile, or those living in urban areas - consistently exhibit lower care-seeking, and consequently are more likely to go untreated ([8]).

Should a person seek healthcare, their probability of receiving treatment depends on the health system’s capabilities. This, in turn, depends on the availability of healthcare workers, equipment, and consumables (medicines and diagnostic tools). Healthcare worker capabilities - such as diagnostic accuracy - also influence treatment. The TLO model was calibrated based on the services delivered, with elasticity in the time required for delivery of each appointment. For this analysis, we assume no restrictions on healthcare worker time; we do assume limitations on consumable availability, with an average availability of 49%, varying by item across facility types, districts, and months ([15]).

In Malawi, treatment and prevention programmes for HIV, TB, and malaria (HTM) are funded via disease-specific (“vertical”) programmes, and are thus subject to different constraints on resources compared to other diseases ([13]). In this analysis, we assume that coverage and consumable constraints for these programmes remain constant over time.

### 3.4 Analysis

We use this model to simulate the changes in population size ([6]), disease burdens, causes of morbidity and need for healthcare in Malawi between 2020 and 2070 (inclusive). We selected this time horizon to capture the progression of diseases associated with aging, which are expected to become more prominent as the population grows older over the coming decades. The causes of morbidity explicitly modelled include: maternal and neonatal conditions, congenital disabilities, acute lower respiratory infections (ALRI), childhood diarrhoea, AIDS, malaria, measles, tuberculosis, schistosomiasis, heart disease, kidney disease, diabetes, stroke, bladder cancer, breast cancer, oesophageal cancer, prostate cancer, depression and self-harm, epilepsy, chronic obstructive pulmonary disease, road traffic injuries (RTI), and lower back pain. For details on how these are modelled, see the model documentation at www.tlomodel.org.

In the main analysis, we assume that the current trends in risk factors, access to treatment, and healthcare-seeking behaviour are sustained (the “Status Quo” scenario). Then, given the inherent uncertainty associated with projections, we consider a number of alternative scenarios which vary in terms of the population’s healthcare-seeking behaviours, the healthcare system functioning (including accuracy of diagnostics and referrals and healthcare worker competence, but not including healthcare worker time), the funding of specific vertical programmes, and consumable availability (SI Taable 1). These sensitivity analyses are detailed in the Supplementary Information.

The model was run with an initial simulated population size of 100,000 individuals (scaled to match Malawi’s actual population size) between 2010 and 2070, with any changes to parameters in the sensitivity analysis implemented in 2020. Our primary outputs were disability-adjusted life years (DALYs), deaths, and healthcare worker time. We also calculated age-standardised disease prevalence (SI).

### 3.5 Ethical Approval

The Thanzi La Onse project received ethical approval from the College of Medicine Malawi Research Ethics Committee (COMREC, P.10/19/2820) for the use of publicly accessible and anonymised secondary data. No data were used requiring individual informed consent.

## 4 Results

### 4.1 Trends in population size and structure

We project that Malawi’s population will increase from around 19.5 million in 2020 to 54.2 million by 2070. Life expectancy (LE, Figure 1) increases from 52 for men and 53 years for women to 66 and 72 years, respectively. The large and persistent discrepancy in life expectancy between men and women is driven in this scenario by a higher number of deaths in men due to road traffic injuries, COPD, and cardiometabolic conditions (SI Figure 7). The life expectancies we project are lower than the World Population Projections from 2024 (WPP, [32]), indicating that to match these projections, an improvement in health (either through lifestyle changes or healthcare access/provision) would be required.

**Figure 1:**
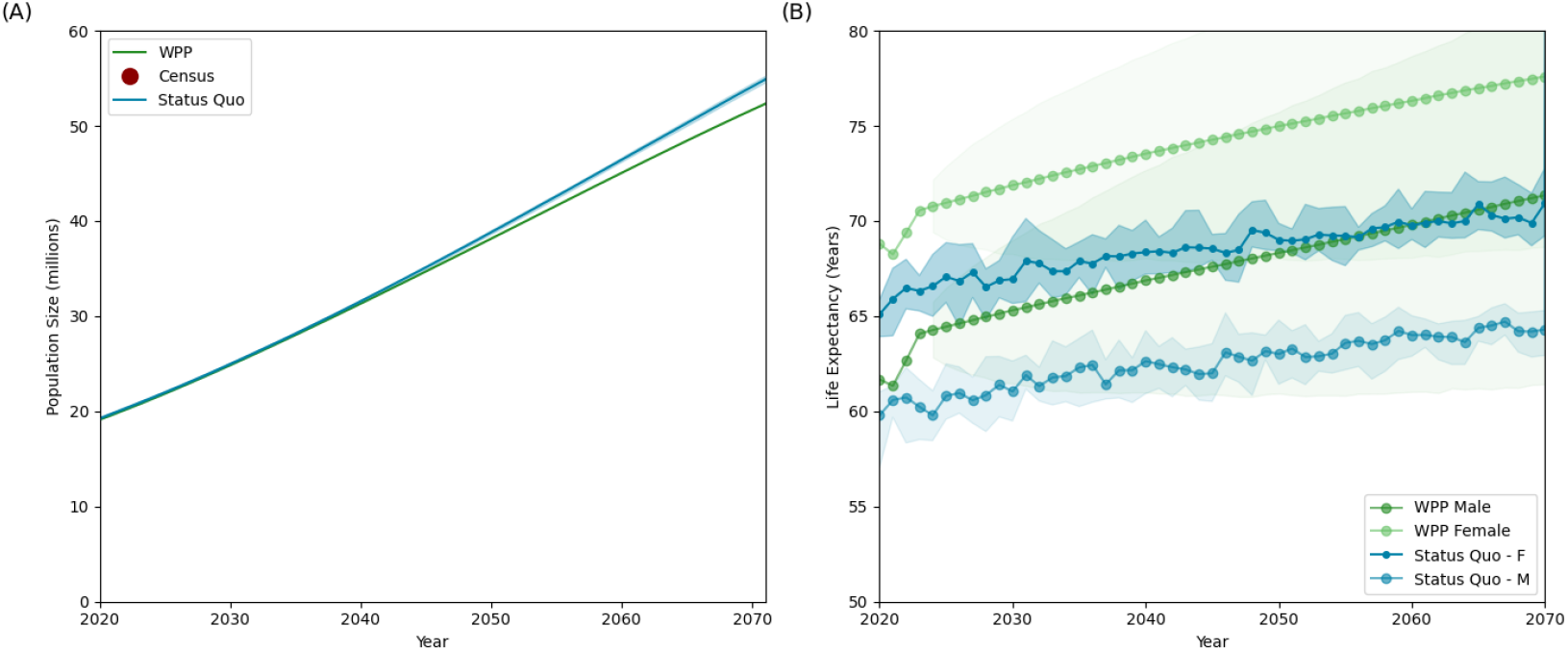
(A) Projected population size (B) Projected life expectancy in the scenario. WPP are the World Population Projections from 2024.

In 2020, Malawi’s population pyramid takes the typical shape of that of an expanding population, with a broad base reflecting high fertility rates and a low proportion of the population over the age of 50. By 2070, we project there will be an expansion of the upper levels of the pyramid, driven by reduced mortality and increased life expectancy.

This is reflected in the median age statistic, which in 2020 is 17 years, and is projected to increase to 25 by 2070. Compared to the World Population Projections (WPP), the TLO model predicts more births and a younger age structure, but under alternative assumption whereby access to and provison of healthcare is greatly increased (the “Maximal Healthcare Provision” scenario in our sensitivity analysis) much more closely matches the WPP projections (SI Figure 9(C)).

### 4.2 Causes of death and ill-health

By 2070, infectious diseases such as AIDS, TB, ALRI, and malaria become less prominent causes of DALYs (Figure 3) AIDS accounts for 0.78% of total DALYs in 2070 compared to 12.8% in 2020, TB is 4.7 vs 6.1%, ALRI 10.6 vs 13.5%, and malaria 2.2 vs 2.9%. This coincides with an increase in the share of DALYs caused by cancers, cardiometabolic diseases, and depression and self-harm. Indeed, in 2020, across all scenarios, 21.8% of DALYs are caused by these conditions; by 2070, 36.7% of DALYs are. DALYs due to AIDS also decrease 6.5-fold, due to assumed increases in treatment accessibility and prevention programmes limiting the spread of HIV.

**Figure 2:**
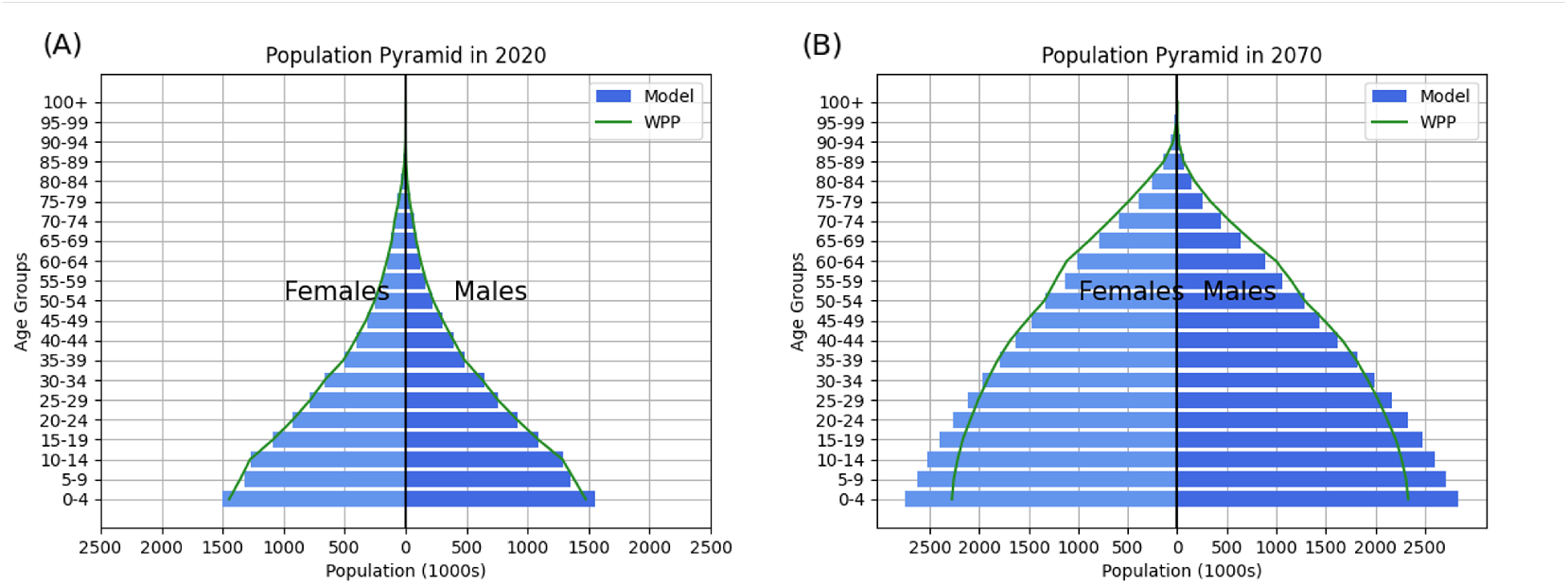
Comparison of population size and age structure compared with the 2024 WPP for (A) 2020 and (B) 2070

**Figure 3:**
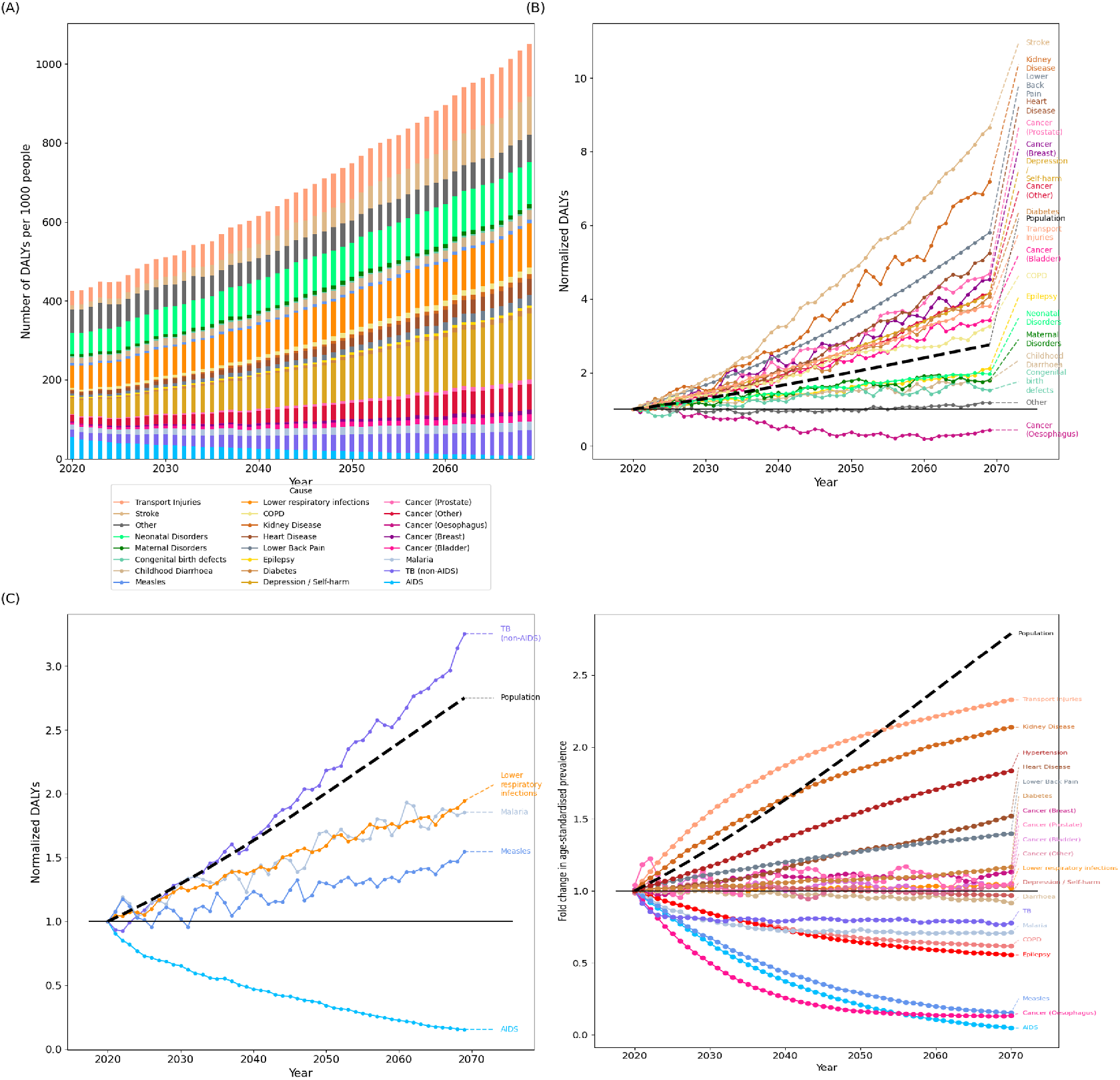
(A) Cause-specific DALYs per 1,000 population, (B) fold change in DALYs for non-communicable diseases each year compared to 2020, and (C) fold change in DALYs for communicable diseases each year compared to 2020. The black lines in (B) and (C) shows the relative growth in population compared to 2020. (D) Age-standardised prevalence of diseases per 1,000 in the population in 2070. The age-standardisation was conducted using the 2020 population as the standard population.

We project an increase in the age-standardised prevalence of many NCDs by 2070 (Figure 11), most notable cardiometabolic diseases such as kidney disease, stroke, and diabetes. The large increase in the prevalence of road traffic injuries is related to the increased proportion of the future population living in urban areas, a major risk factor for RTI in the model.

Notably, these trends persist even in an alternative scenario whereby risk factors for developing NCDs become gradually less common (the “Improving Lifestyle Factors” scenarios (SI Figure 11)).

Ongoing improvements in the HTM programmes have meant that the prevalence of HTM will tend to decrease over time (SI Figure 11). However, even greater reductions are possible with targeted scale-up of these programmes (see the HTM Scale Up scenario in the sensitivity analysis, which represents an increase in the coverage of treatment and prevention programmes for HIV, TB and Malaria). However, only under the “Maximal Healthcare Provision” scenario are total DALYs substantially reduced overall (SI Figure 10).

### 4.3 Need for the healthcare workforce

Long-term projections show that,the time required to deliver care will increase for all cadres of healthcare. The greatest proportionate increase is observed for the mental health cadre (Figure 4). This is despite the relatively stable prevalence of depression in the population (Figure 10D). However, the ageing population drives an increase in the number of people in need of such care, even though the risk of onset and overall proportion of the population with those conditions does not increase. Additionally, there were very few mental health staff working in 2020, so there was a small initial denominator for the relative change. The most in-demand appointments included those expected to increase with a growing population (Figure 5), such as those related to HTM prevention and road traffic injuries.

**Figure 4:**
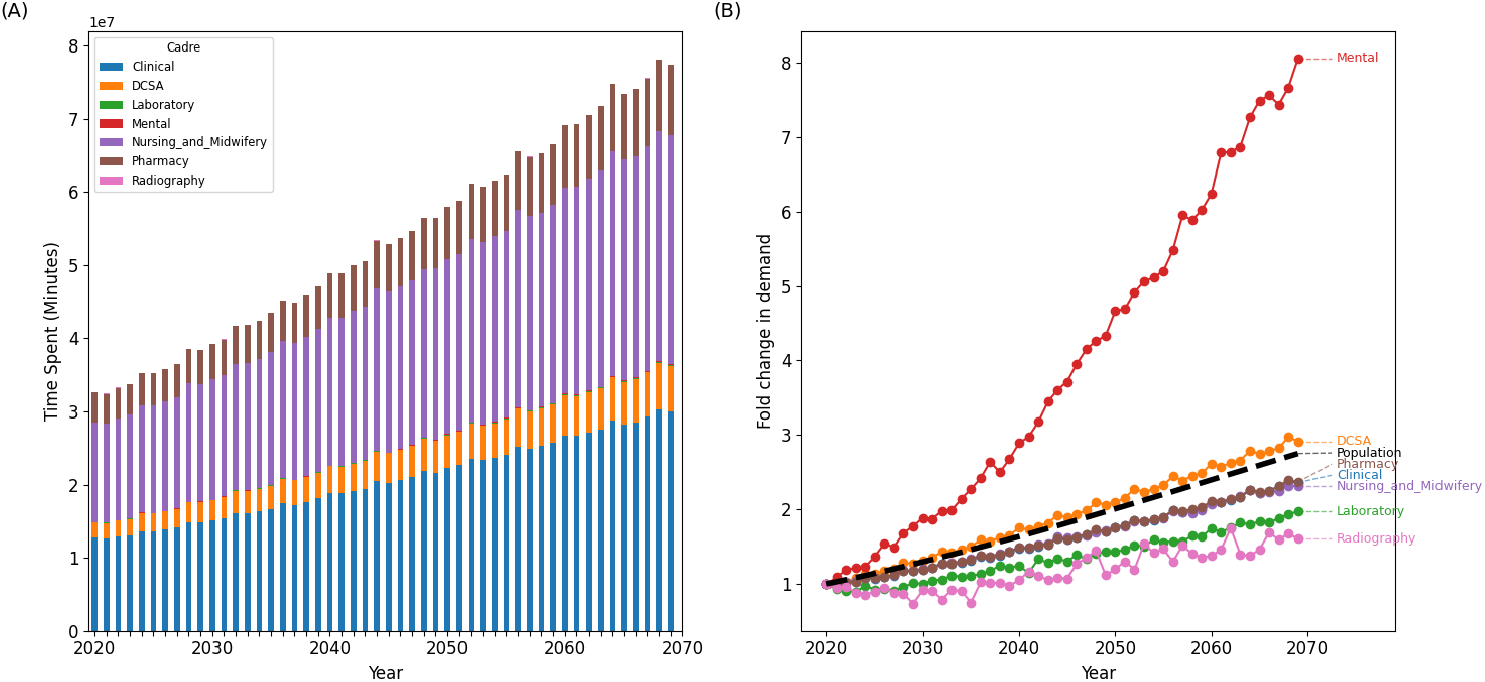
(A) Time spent (in minutes) by each cadre between 2020 and 2070 in the population. (B) Fold change in 2070 compared to 2020. The black line in (B) shows the relative population growth each year compared to 2020.

**Figure 5:**
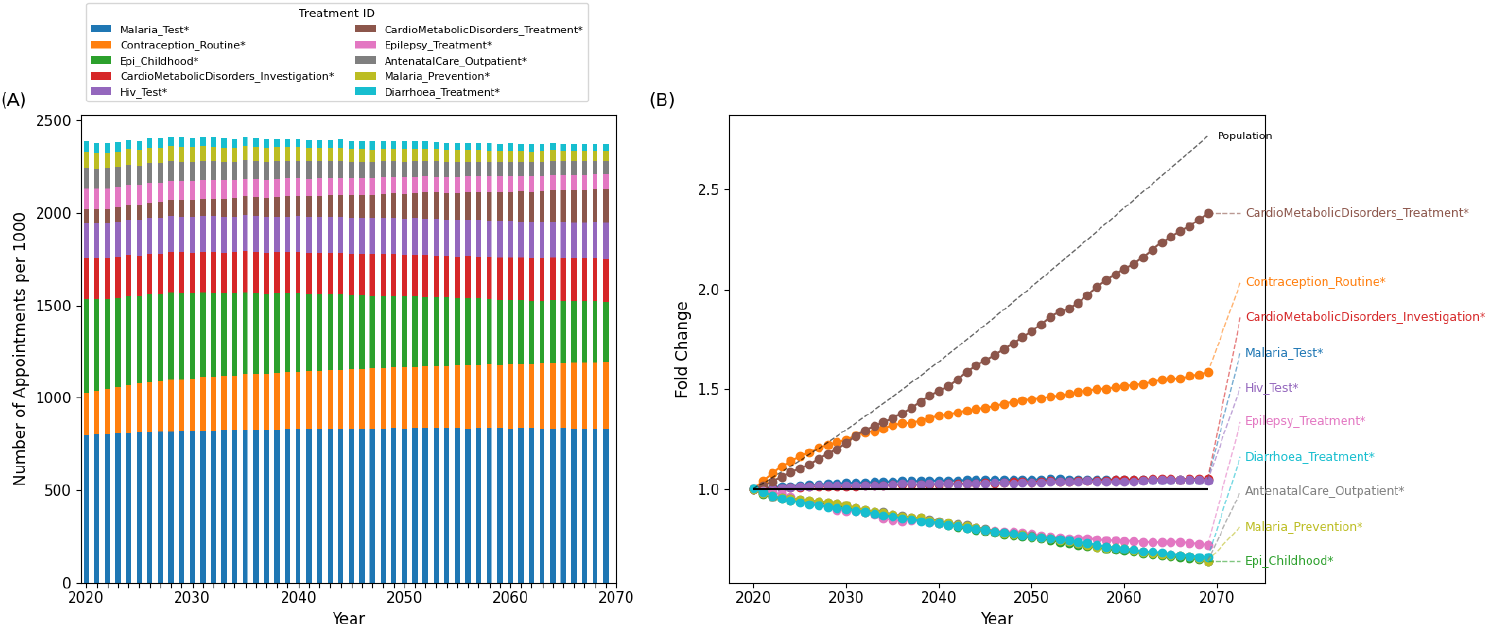
(A) Number of interactions per 1,000 population of the top-ten most needed interaction types between 2020 and 2070. (B) Fold change in 2070 compared to 2020.

Under the scenario of increased healthcare service access and provision (the “Maximal Healthcare Provision” scenario in the sensitivity analyses), the overall demand for the HCW time increases (SI Figure 12), This is driven by increased healthcare-seeking behaviour and arises despite the lower actual disease burden that the population would come to incur.

## 5 Discussion

Here we present a set of combined epidemiological, demographic, and health-care demand projections for Malawi. We find that there will be a simultaneous growth and ageing of the population by 2070, with a concomitant rise in the prevalence of NCDs. Both of these factors will result in a significant increase in healthcare demand for treatments related to cancers, depression, and cardiometabolic disorders, with an associated increase in demand for the clinical and mental health cadres.

By 2070, we project Malawi’s population to increase to around 54 million people (Figure 8A), and despite larger numbers in older age groups, its young population structure remains (by 2070, the median age remains 25 years), in broad agreement with other projections [32]. Though our “Status Quo” scenario as presented in the main text projects more births and a lower life expectancy than the WPP (Figures 8 - 2), our “Maximal Healthcare Provision” scenario in the sensitivity analysis aligns well (SI Figure 8 - 9). This indicates that for our baseline projections to match those of the WPP, a change in lifestyle factors and/or better access to healthcare services (including contraception) is necessary. Importantly in this scenario, the availability of contraception means that population growth decreases, analogous to when contraception is used in family planning campaigns ([6])

**Figure 6:**
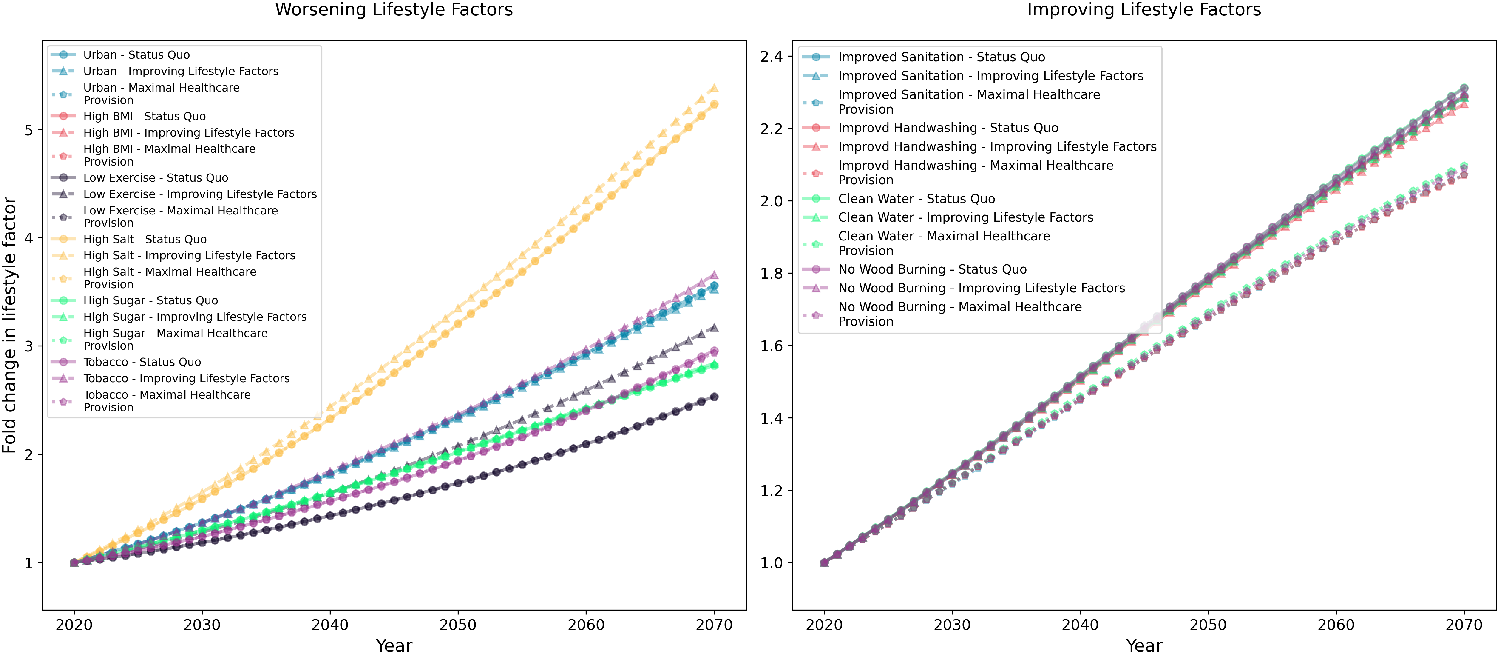
Change in lifestyle factors over time in the Status Quo, Worsening Lifestyle Factors, and Improving Lifestyle Factors scenarios. (A) The factors examined in the Worsening Lifestyle Factors scenario and (B) those under the Improving Lifestyle Factors scenario. The patterns in the HTM Scale-Up and Maximal Healthcare Provision reflect those of the Status Quo.

**Figure 7:**
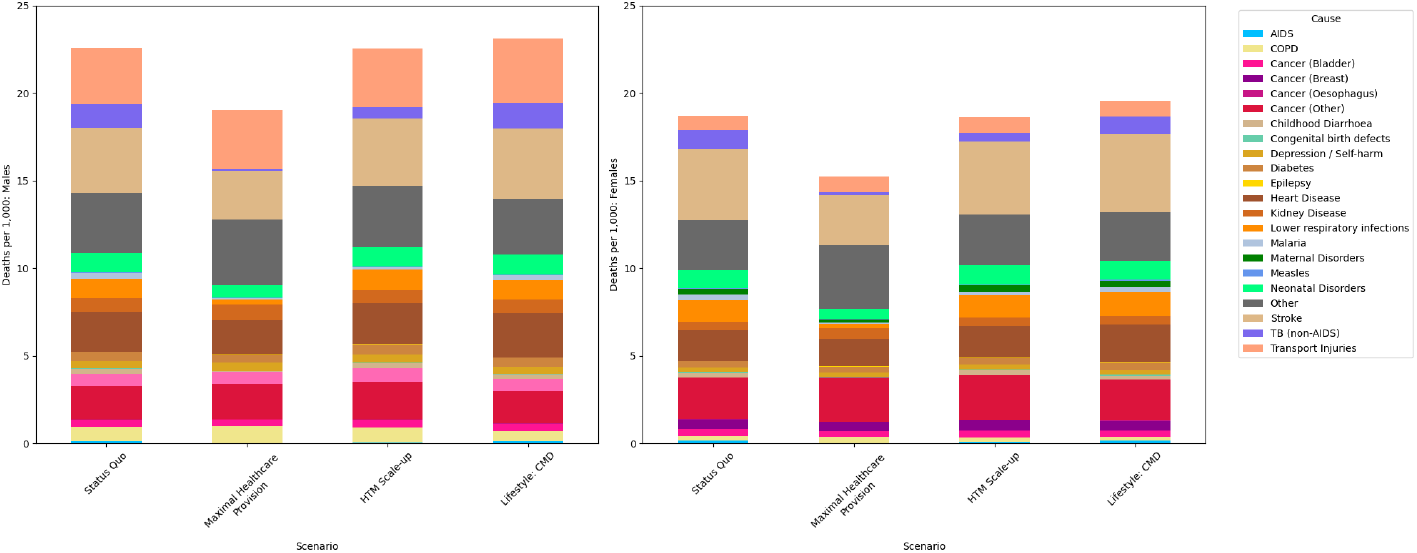
Cause of death per 1,000 people amongst the (A) male and (B) female population in 2070.

**Figure 8:**
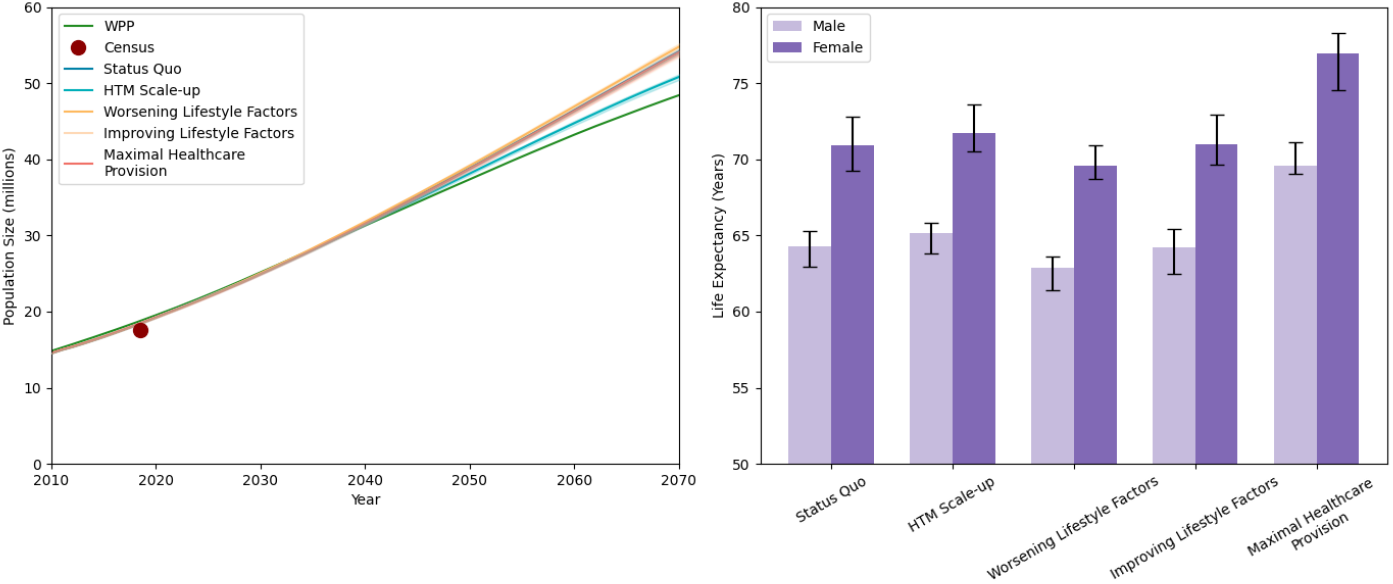
(A) Projected population size. (B) Projected life expectancy across all modelled scenarios.

Alongside any differences in assumptions about healthcare access and lifestyle factors, differences between the WPP and the TLO model could be due to differences in methodology. The WPP is based on statistical extrapolations of observed historical trends, whilst the TLO estimates are based on mechanistically modelling diseases, comorbidities, and access to healthcare.

Our sensitivity analysis indicates that the largest gains in LE are probably driven by reduced mortality due to infectious diseases such as TB, HIV, and ALRI (Figure 8B). This is most prominent in the scenarios that “target” investment in the vertical HTM programmes guided by the National Strategic Plan for HIV and National Malaria Control Plan, highlighting the importance of investment in these programmes.

Recent decades have seen an increase in the burden of NCDs in Malawi, a trend projected to continue: by 2070, we project an estimated 37% increase in the proportion of deaths and DALYs due to NCD under the Status Quo scenario (SI Figure 13). However, despite the historic rise in the prevalence of NCDs, access to related healthcare is limited. Services have been historically underfunded [11, 24] and lacked sufficient equipment and personnel [14, 7, 29], resulting in low quality of treatment, particularly in terms of NCD prevention ([11]). The increase in NCDs occurs in the context of still-high prevalences of infectious diseases, resulting in a double burden of disease. Some infectious diseases are risk factors for NCDs (for example, HIV infection is a risk factor for depression), so continued burdens of these infectious diseases exacerbates the age-related increase in prevalence. With inadequate allocation of resources to the prevention, screening, and treatment of these conditions [24], coupled with a lack of knowledge in the population about their risk [7, 24], means that the increased burden of NCDs that we project will only strain the healthcare system more.

The most significant increase in demand for healthcare worker time was for the mental, clinical, pharmacy, and nursing and midwifery cadres. There was a significant increase in demand for the mental health cadre, related to the approximately four-fold increase in number of DALYs caused by depression observed across all scenarios (SI Figure 10(B)). The increase in mental health conditions may be related to the high prevalence of other risk factors, such as HIV and ageing (the latter accounting for the fact that, despite the increase in DALYs, the age-standardised prevalence in the population remains relatively constant over time (SI Figure 11). However, for appointment types, those related to HTM prevention and RTIs were the most needed. HTM prevention schemes scale with population growth, as do road traffic accidents, so this is expected. The increase in urbanicity over time also contributes to the increasing prevalence of RTIs.

The complexity of the model, and the long-term nature of the projections, necessitate certain simplifying assumptions. We assume, for example, that no new treatments or medical technologies become available in Malawi during our time horizon. This assumption will have a particular impact on the estimates of ill-health caused by NCDs, which currently have few treatment options. Similarly, for RTIs, we assume that there is no change in road safety between 2020 and 2070, though there is a national strategy for its improvement ([26]).

The fertility schedules are dependent on the availability of contraception, which is limited in all scenarios except the “Maximal Healthcare Provision”. The TLO model does not account for private healthcare, which makes up a small but potentially increasing share of all healthcare in Malawi [25].

Across all scenarios, in order to find the total need for healthcare worker time, no constraints were placed on the total time that will actually be available.. In the (likely) case that need for healthcare time outstrips what is available, then the total healthcare delivered will be less than shown here and disease burdens would therefore likely become greater as a consequence. [8].

We also assume that the productivity and competence of healthcare workers remains constant over time, which could under-estimate service delivery also.

Malawi is projected to undergo simultaneous demographic and epidemiological transitions, both of which will impact the demand for healthcare services in the medium and long-term. The projections uniquely leverage the TLO model, which mechanistically accounts for interactions across the whole health system and shows how this generated future need for healthcare services. Overall, as the burden shifts from infectious and childhood diseases toward NCDs and age-related conditions, the healthcare system must adapt to meet those changing demands.

## Data Availability

The Thanzi La Onse model is open source and available for review and usage at https://github.com/UCL/TLOmodel. The files used in this analysis are found at:https://github.com/UCL/TLOmodel/tree/rmw/long_term_projections

## 6 Supplementary Information

### 6.1 Scenarios for projecting long-term trends

Our main scenario (Status Quo) assumes that all parameters governing the onset of disease, healthcare-seeking behaviour and healthcare system function remain at current levels (as detailed in the main text).

Our second scenario (“HTM Scale-up”) scales the HIV, tuberculosis (TB), and malaria (together, HTM) vertical programmes to meet the 95-95-95 target set out by the 2030 Global Aids Strategy [18] and National Malaria Control Plan [12] (hereafeter referred to as “Target” levels), with all other aspects of the healthcare system and risk factors remaining unchanged from the status quo.

The third scenario (“Worsening Lifestyle Factors”) assumed accelerates changes in lifestyle factors that, in turn, influence the risk of developing a range of diseases, but were in particular chosen for their influence on developing NCDs. These factors are the three-month risk of developing higher BMI, switching to a high-salt diet if an urban resident, switching to a high-sugar diet, reducing exercise (if male), initiating tobacco use (if male), and consuming excess alcohol (if male) (Table 1). The baseline probability of exposure to each risk factor is increased by 50%; if there is a relative risk due to other factors (such as age, if living in an urban area, etc.), or a time-dependent change in this risk factor, these are enacted on this modified baseline.

**Table 1:** Assumptions around population behaviour and healthcare system parameters across scenarios. Details of the “Lifestyle” risk factors can be found at www.tlomodel.org. The “Default” levels are those found in the Status Quo scenario. “Target” indicates that HTM programmes are scaled to the Global AIDS Strategy and National Malaria Control Plan targets. “Perfect” healthcare seeking means that all those who develop symptoms receive care; default healthcare seeking is determined by factors including severity of symptoms, wealth quintile, and age.

| Scenario | Healthcare System Functioning | Consumable Constraints | Vertical Programme Funding (HTM) | Healthcare Seeking Behaviour | Lifestyle Factors |
| --- | --- | --- | --- | --- | --- |
| Status Quo | Default | Default | Default | Default | Default |
| Maximal Healthcare Provision | Maximum | None | Target | Perfect | Default |
| HTM Scale-up | Default | Default | Target | Default | Default |
| Worsening Lifestyle Factors | Default | Default | Default | Default | **50% increased probability per 3 months of:**<br>• Developing higher BMI (rural males 15–29, non-tobacco users, low wealth/sugar): 0.00075<br>• High salt diet (urban residents): 0.0045<br>• High sugar intake: 0.00015<br>• Reduced exercise (rural males): 0.0015<br>• Starting tobacco use (males 15–20, low wealth): 0.0006<br>• Excess alcohol consumption (males): 0.0045 |
| Improving Lifestyle Factors | Default | Default | Default | Default | **50% increased probability per 3 months of:**<br>• Eliminating wood-burning stove: 0.0015<br>• Gaining clean drinking water access: 0.0015<br>• Improving sanitation facilities: 0.0015<br>• Gaining handwashing facilities: 0.0015 |

Such change in risk factors could increase future health demand, particularly when considered alongside the anticipated changes in demography. However, these risk factors are linked to NCDs, some - such as lack of sanitation facilities, or access to a wood burning stove - also influence other areas of health. While the model accounts for the possibility of losing these risk factors (e.g., developing a lower BMI), the lower probabilities associated with such transitions lead to a growing tendency toward these risk factors over time.

In the fourth scenario (“Improving Lifestyle Factors”), we only increase the probability of factors that reduce the risk of disease (the three-month probability of switching from a wood-burning stove, gaining access to clean water, having improved sanitation, and gaining access to handwashing facilities). For these parameters, we assume that once access is gained, it is retained permanently (i.e. there is no reversal of trend).

An increase of 50% in lifestyle risk factors reflects a plausible upper-bound estimate of potential behavioural and economic shifts. The rate of urbanisation is increasing ([30]), which is a risk factor for obesity ([2]), high salt intake ([16]), and smoking ([3]). Changes in diet, including increasing salt and sugar intake (again, both linked with higher BMI) are increasingly prevalent ([9, 33]). The “positive” lifestyle changes an upper bound if the continuing improvement in access to sanitation facilities continues ([31]).

Our final scenario, the “Maximal Healthcare Provision”, aims to understand how population health, and consequent healthcare demands, will change over time if frailties in the healthcare system are removed. It imposes no limitations on the consumable, and that the accuracy of diagnoses and referrals is improved (in a disease-dependent manner). We also assume that there is perfect healthcare-seeking behaviour within the population. Additionally, the vertically-funded HTM programmes are scaled up to meet the aforementioned targets.

## 6.2 Supplementary Results

### 6.3 Sensitivity Analysis

The broad trends in population size and life expectancy are replicated across other scenarios (Figure 8). The population size in 2070 varies between 50.8 and 54.8 million (in the “Maxmimum Healthcare Provision” and “Vertical Programme Funding” scenarios, respectively). Unsurprisingly, highest LE gains are achieved under the “Maximal Healthcare Provision” Scenario (reaching 70 and 78 years for men and women), primarily because of the drop in prevalence of infectious diseases of childhood. This is also reflected in the HTM Scale-Up scenario. Perhaps counter-intuitively, this also resulted in the lowest population size (50.8 million), related to the lower fertility rates driven by the availability of contraceptives ([6]).

Interestingly, our projections under “Maximal Healthcare Provision” scenario most closely resembled those of the World Population Projections (Figure 9(C)), indicating that to achieve the projections laid out by the WPP, an increase in health service availability or utilisation would be required.

**Figure 9:**
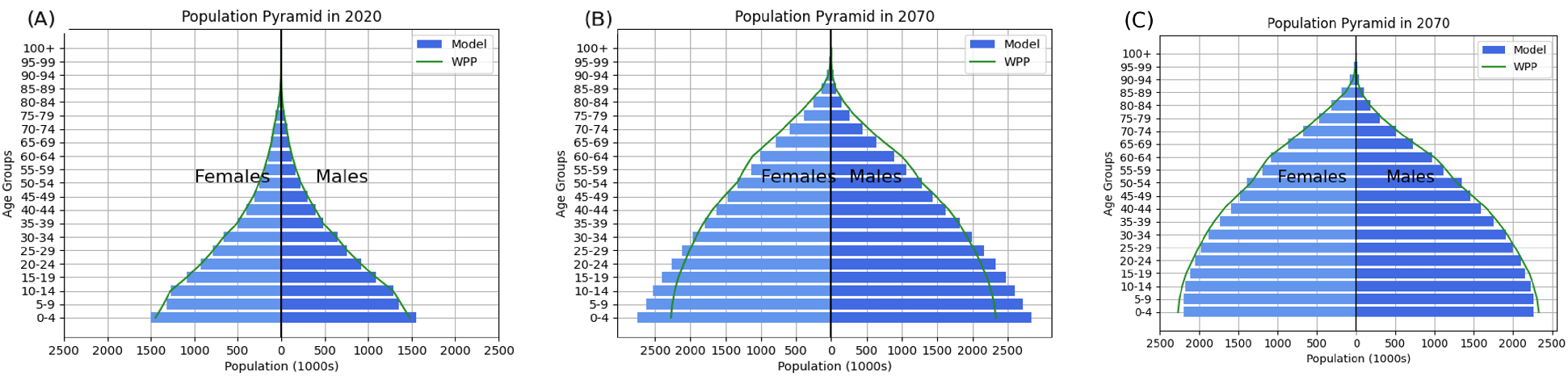
Comparison of population size and age structure under the “Status Quo” scenario for (A) 2020 and (B) 2070. (C) Shows the population age structure under the “Maximal Healthcare Provision” scenario.

**Figure 10:**
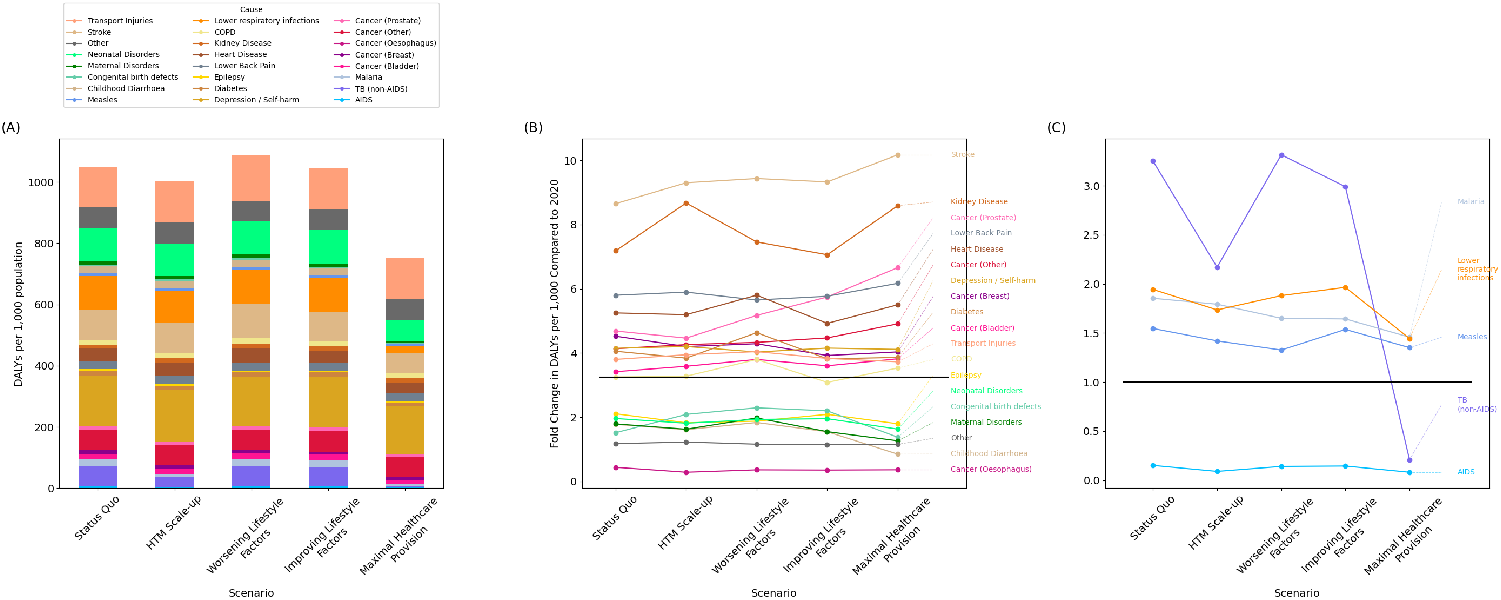
(A) DALYs per 1,000 in 2070 and (B) relative change in DALYs per 1,000 by cause for 2070 for each scenario.

These broad trends in DALYs are replicated across our scenarios. However, the “Maximal Healthcare Provision Scenario” saw larger increase in DALYs caused by NCDs (48%). The increase in NCD prevalence in the “Maximal Healthcare Provision” scenario was higher than the increase observed for the “Worsening Lifestyle Factors”. Indeed, the highest increase in prevalence of NCDs was observed under the “Maximal Healthcare Provision” due to the lowered mortality from diseases of childhood (ALRI, diarrhoea) or other infectious diseases (TB, malaria, AIDS). The increased prevalence could also be related to the improved healthcare-seeking behaviour and treatment, which may cause cancers to be diagnosed earlier and therefore persist longer in the population. Indeed, for across all cancers, there are fewer years of life lost due to premature mortality (YLLs) and years of healthy life lost due to disability (YLDs) in the “Maximal Healthcare Provision” scenario compared to the others (SI Figure 14).

**Figure 11:**
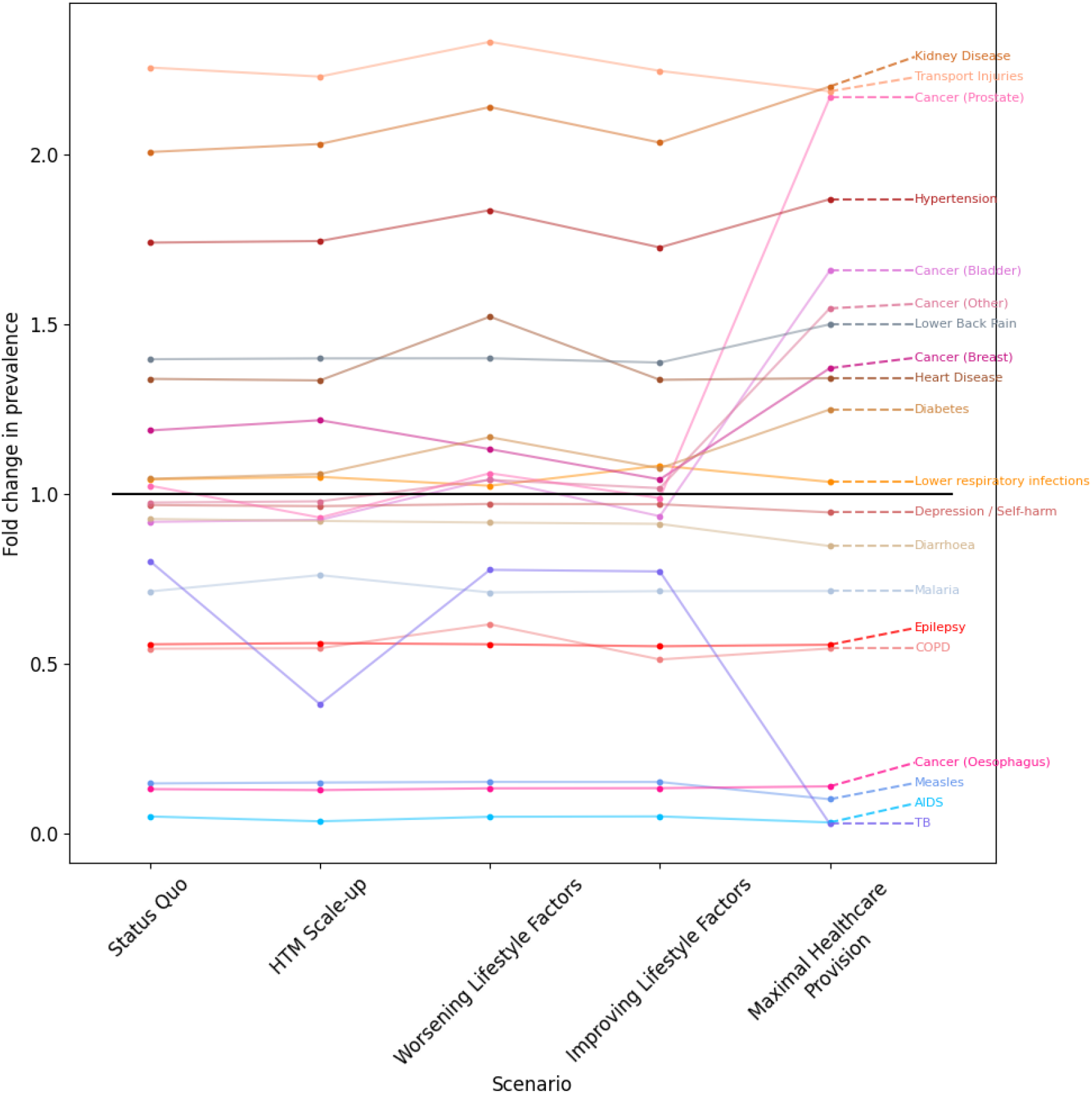
Fold change in the age-standardised prevalence of diseases per 1,000 in the population in 2070 compared to 2020. The black line indicates the fold change in population growth.

**Figure 12:**
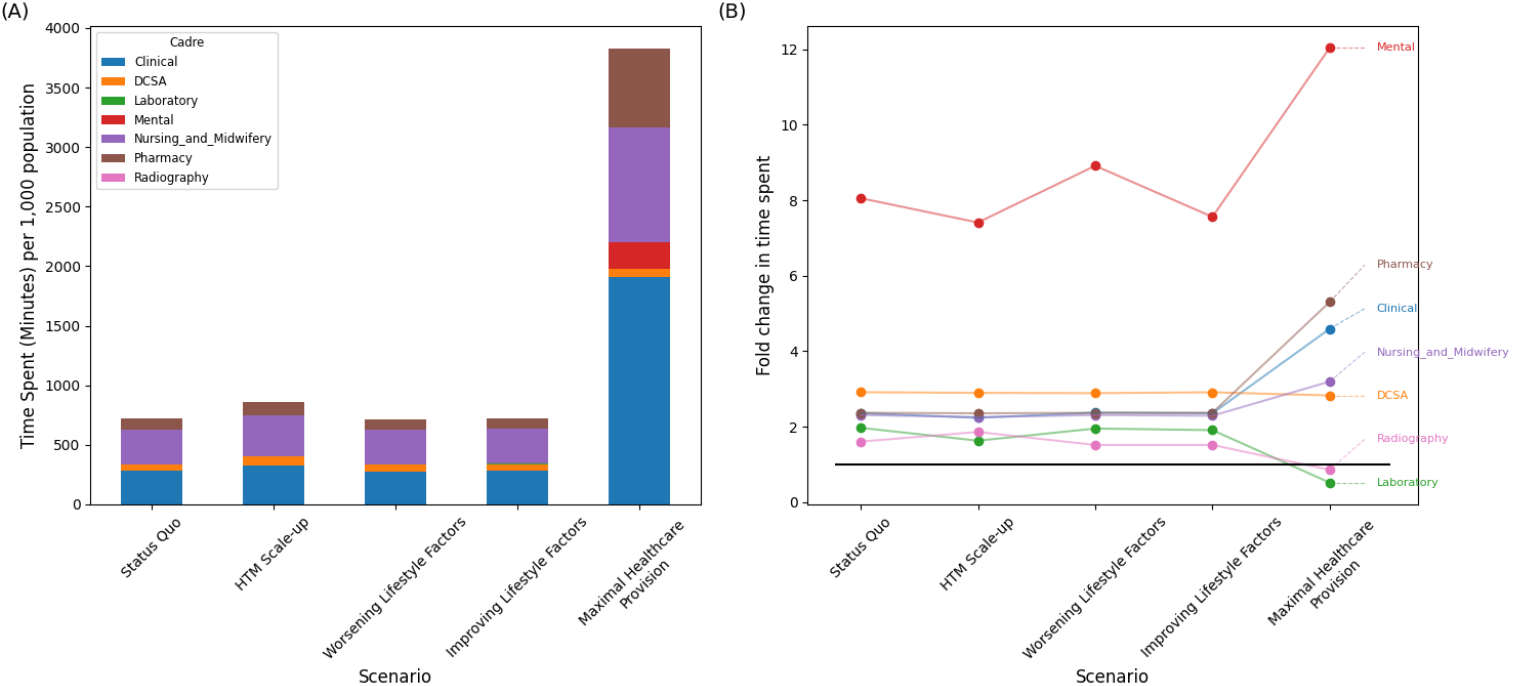
(A) Time spent (in minutes) by each cadre between 2020 and 2070 per 1,000 in the population. (B) Fold change in 2070 compared to 2020.

**Figure 13:**
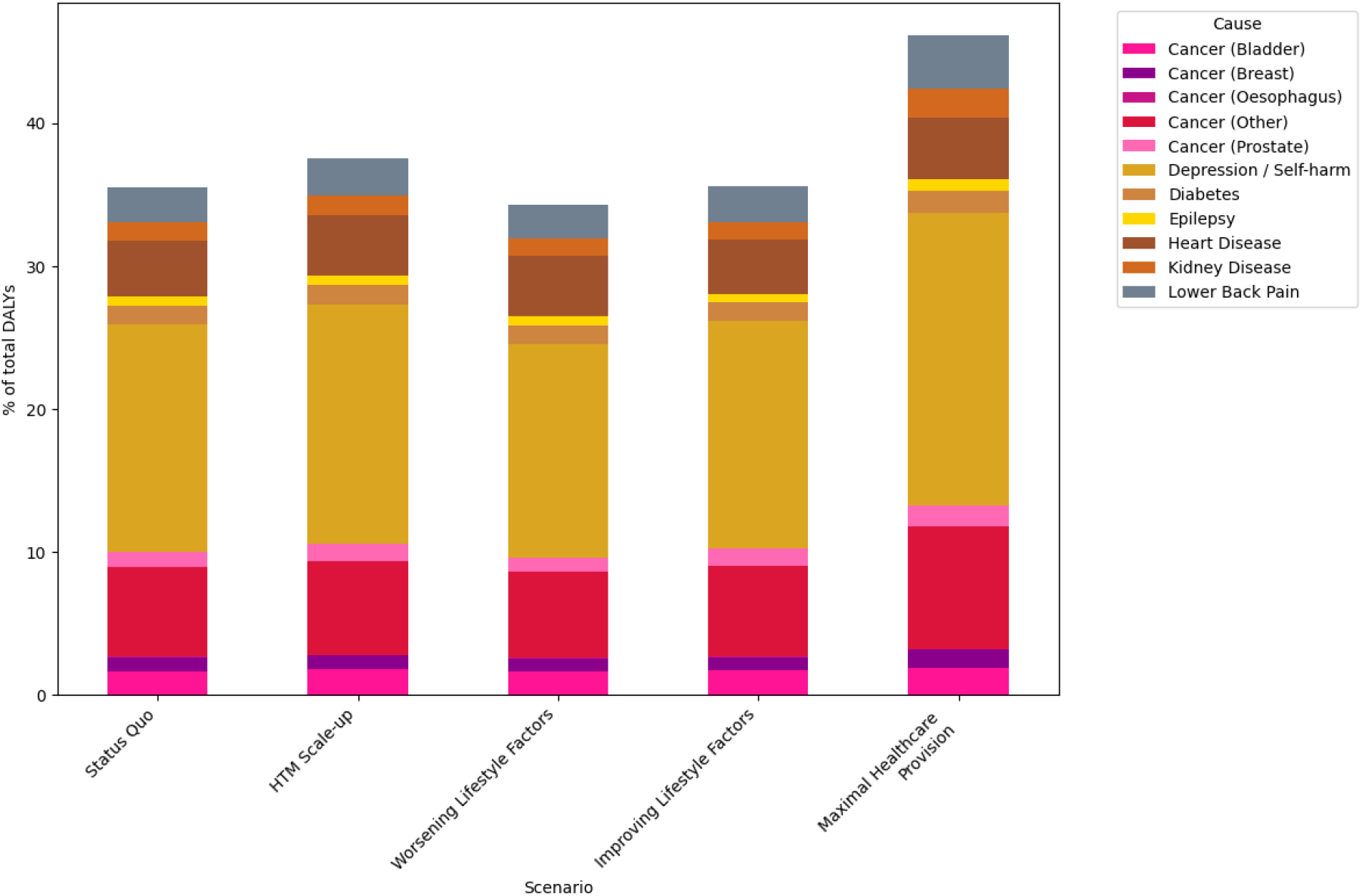
Projected share of Disability-Adjusted Life Years (DALYs) attributable to non-communicable diseases (NCDs) in 2070 by scenario.

**Figure 14:**
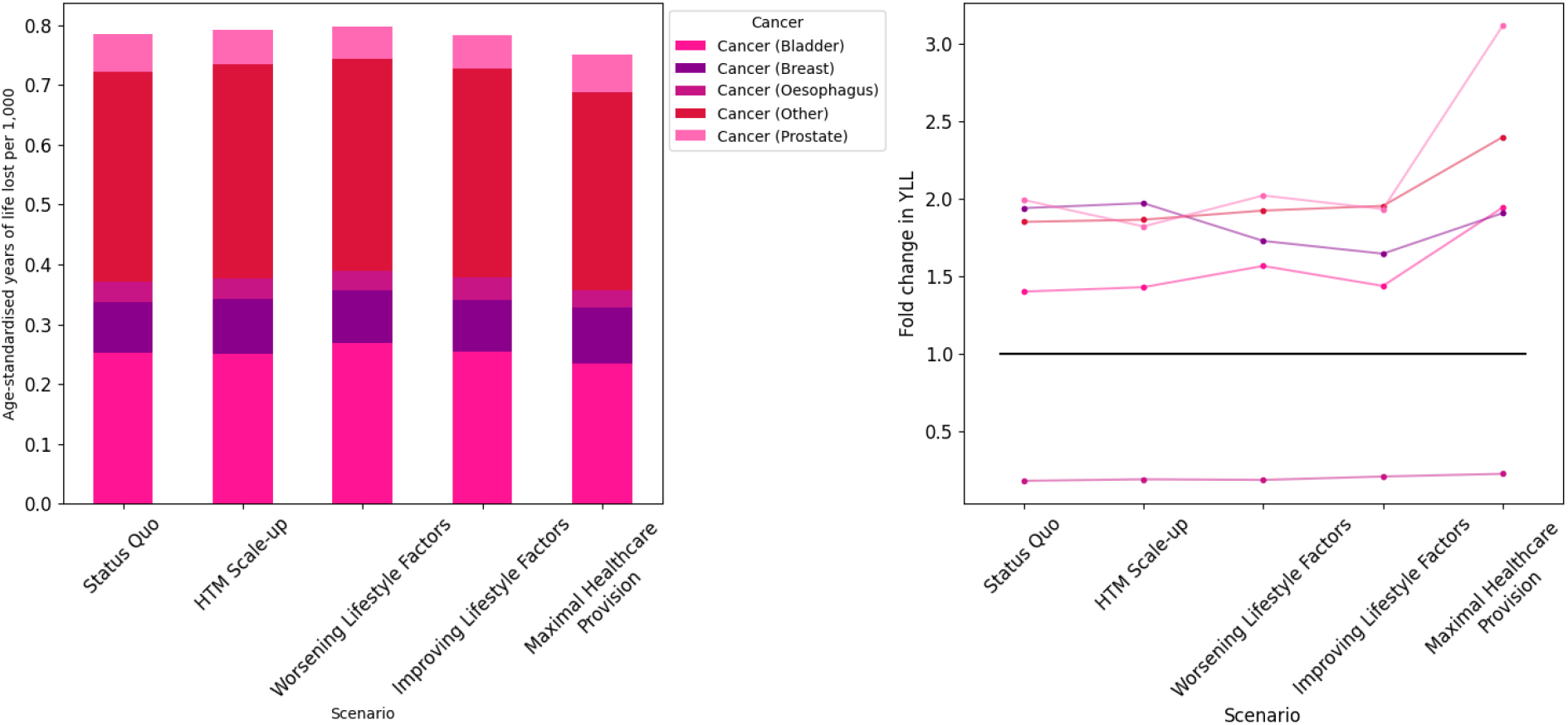
(A) Years of life lost (YLL) due to cancers in 2070. (B) Fold change in YLL between 2020 and 2070.

